# Efficacy and Safety of Viable Selective Germline Genome Edited Pigs Skin Xenotransplants in Patients with Thermal Burns

**DOI:** 10.1101/2021.12.30.21267448

**Authors:** Lijin Zou, Youlai Zhang, Ying He, Hui Yu, Fan Yang, Jian Huang, Fangjuan Li, Qianglong Sheng, Yeming Zhang, Yajun Li, Fei Chen, Guangqian Zhou, Xuenong Zou, Qingqing Wang, Hong-ye Zhou, Ningyan Hu, Yuanlin Zeng, Hong-jiang Wei, Yin Yu, Gang Wang

## Abstract

**Background:** Rapid closure of open wound, either temporarily or perpetually, is recognized as the standard of care in patients with thermal burns. Human cadaveric allograft and simple genetically modified porcine xenografts are not able to provide enough durable time for extensively burned patients. A selective germline genome edited pig (SGGEP) skin xenograft, Xeno X skin, would be a valuable candidate to the clinical options.

**Methods:** In an ongoing investigator-initiated clinical trial in patients with thermal burns, the efficacy and safety of cryopreserved Xeno X skin grafts of SGGEP for burned patients were evaluated. Each patient received surgical grafting with a skin xenotransplant and wild type pig extracellular matrix (wpECM) in a side-by-side manner for in-situ comparison.

The primary outcome measures of xeno-skin grafts included Xeno X skin safety and tolerability, as well as the quality and duration of temporary barrier function yielded by Xeno X skin grafts (as determined by Baux score). Seven parameters included in the analysis were vascularization, pigmentation, thickness, relief, pliability, surface area and the overall opinion, with each calculated on an independent 0-10 scale.

**Results:** A total of 16 burned patients completed the trial. All the patients tolerated Xeno X skin grafts well and no advent events were observed. In all cases, Xeno X skin grafts were vascularized and fully adherent, they also exhibited better overall outcomes than those of wpECM. Xeno X skin grafts survived for at least 25 days without a need of any immunosuppressive drug, well consistent with our earlier preclinical studies in non-human primates.

**Conclusion:** Xeno X skin grafts of SGGEP did not incur any signs of local and systemic safety issues, and in the meanwhile provided a high quality and long duration of temporary barrier function for burned patients. This is a major milestone in the xenotransplant field, indicating that genome-edited organ xenotransplant has become a clinical reality.

## Introduction

WHO reported there are 11 million burn injuries with roughly 180,000 deaths per year globally (https://www.who.int/en/news-room/fact-sheets/detail/burns).Burns also are an important source of casualty from military openrations^1,2^.

Rapid closure of open wound, either temporarily or perpetually, is always the goal of burn care and is often a life-saving measurement because it suppressed post traumatic fluid loss, infection, and hypermetabolic reactions. Also, it restored the mitochondria energy expenditure^3,4^.

Human cadaveric allograft and porcine xenografts could offer the temporary closure^1,5^,but the survival time for these two materials is round 8 days and 13 days respectively^6-8^. For the patients with extensive and compromised healing ability, these duration time cannot even support a bridge treatment.

Our previous studies have demonstrated that the comprehensive gene edited pig (selective germline genome edited pig, SGGEP) skin xenografts have survived at least 25 days without any immunosuppressor in non-human primates(https://www.biorxiv.org/content/10.1101/2020.01.20.912105v1).

Here we report the safety and efficacy of the cryopreserved SGGEP’s skin grafts (Xeno-X skin) in skin xenotransplant, with wild type pig extracellular matrix (wpECM) in a side-by-side, in-situ comparison without any immunosuppression.

Long immune tolerance(>25days) with no advent events in burned patients were demonstrated bySGGEP’s skin grafts. Although the US Food and Drug Administration (FDA) approved a first-in-human clinical trial phase I (ClinicalTrials.gov Identifier: NCT03695939) to assess the safety and tolerability of a single gene modified pig skin xenotransplant for burned patients in 2018, the current investigation is the first-in-human, prospective cohort investigation in xenotransplantation field with specific data. This is a major milestone in this field and organ xenotransplant has become a clinical reality.

## Methods

### Materials

The SGGEP was established, and the Xeno X skin was manufactured with proprietary processes as we previously described(https://www.biorxiv.org/content/10.1101/2020.01.20.912105v1).We also followed the FDA Guidance for industry: Source Animal, Product, Preclinical, and Clinical Issues Concerning the Use of Xenotransplantation Products in Humans(https://www.fda.gov/media/102126/download).Each Xeno X skin was stored at -70°C to -90°C and went through proper testing before usage.

### Trial oversight

We evaluate the safety and efficacy of Xeno X skin grafts of SGGEP for burned patients. Every patient received surgical grafting with a skin xenotransplant, as well as wild type pig extracellular matrix (wpECM) in a side-by-side way, making in-situ comparison in the absence of immunosuppressor. The Photographic pictures of gross anatomic and pathological results were recorded in indicated time points.

The trial is being conducted in accordance with Good Clinical Practice and the Declaration of Helsinki. The study protocol was approved by the Institutional Review Board of the First Affiliated Hospital of Nanchang University. All participants provided written informed consent before enrollment. Safety was examined by a protocol safety review team weekly and by an independent data and safety monitoring board on a continual basis.

### Patients

Eligible patients were aged >18 years, male or female of non-childbearing potential, who suffered from thermal burns having 3 -32% total body surface area (TBSA). Burn wounds required excision and temporary coverage of wound based on clinical judgement. Each patient has the adequate area of burn wound for Xeno X Skin deployment without a history of allograft or xenograft treatment, and the wound site is not located on face or hands.

Patients were excluded if they are pregnant or lactating women, if they had treatment with immunosuppressive regimens or had a known history of malignancy, allergy to penicillin, aminoglycosides or amphotericin, insulin-dependent diabetes, or other underlying conditions that could undermine their safety. Infected wounds were also excluded.

### Treatment

After excision of the nonviable tissue, Xeno X Skin graft and wpECM were used to their respective, randomly assigned treatment sites in a side-to-side way. Xeno X skin and wpECM were trimmed to fit the wounds, were laid down on the designated treatment sites and secured in place by staples, sutures, or adhesive, based on the investigator’s judgment.

The investigator determined when or whether the patient had need of further autografting after the placement of Xeno X skin and wpECM.

Wounds beyond the study treatment sites were handled according to the First Affiliated Hospital of Nanchang University’s standard of care for that type of injury.

### Efficacy Assessments

The primary endpoint was the efficacy of the Xeno X Skin as illustrated by the quality and duration time of temporary barrier function provided by Xeno X Skin (as quantified by Observer Scar Assessment Scale, POSAS)^9^. Six parameters: vascularization, pigmentation, thickness, relief, pliability, surface area and overall opinion, were gauged.

Each parameter was evaluated on an independent 1-10 scale. It was anticipated that some parameters may not be assessable at all stages of the grafting process and a score of 10 may be recorded.

### Safety Assessments

Safety assessments include monitoring of solicited local or systemic adverse events and serious adverse events, vital signs, laboratory parameters, incidence of wound infection of donor-site for 4 weeks.

### Statistical analysis

Descriptive statistics were calculated using GraphPad Prism Version 9.1.1 (GraphPad software, San Diego, California). Statistical mean is used to present the data..

## Results

### Trail population

Between May 30,2021 and September 30, 2021, sixteen patients were enrolled (11 males, 5 females; median age, 32 years, median %TBSA, 8 (Fig. 1 and Table 1). By week 4, one patient was lost to follow-up. The mean treatment-site area in both groups was below 200cm^2^.

**Figure 1.**
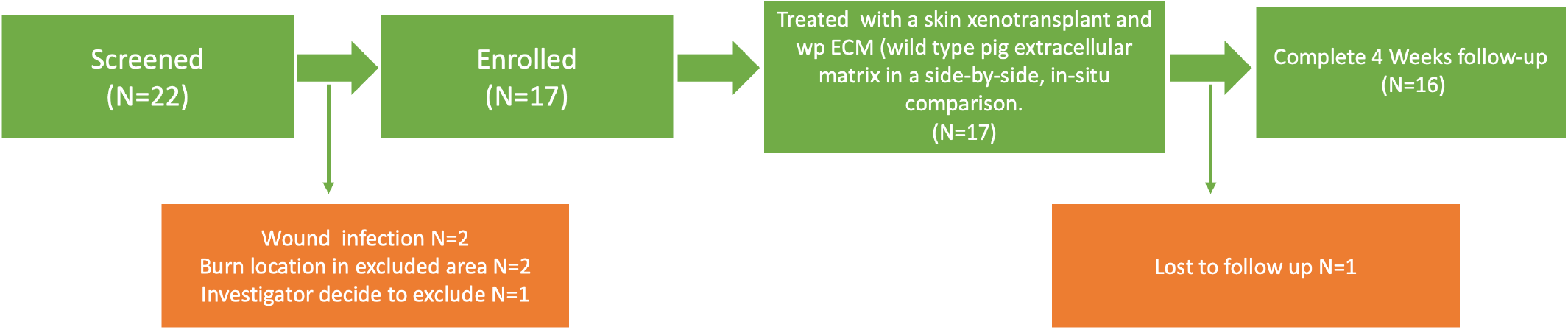
Participant recruitment, and follow-up in the Investigator Initiated clinical trial of skin xenotransplantation for burned patients with the selective germline genome edited pig.

**Table 1.**
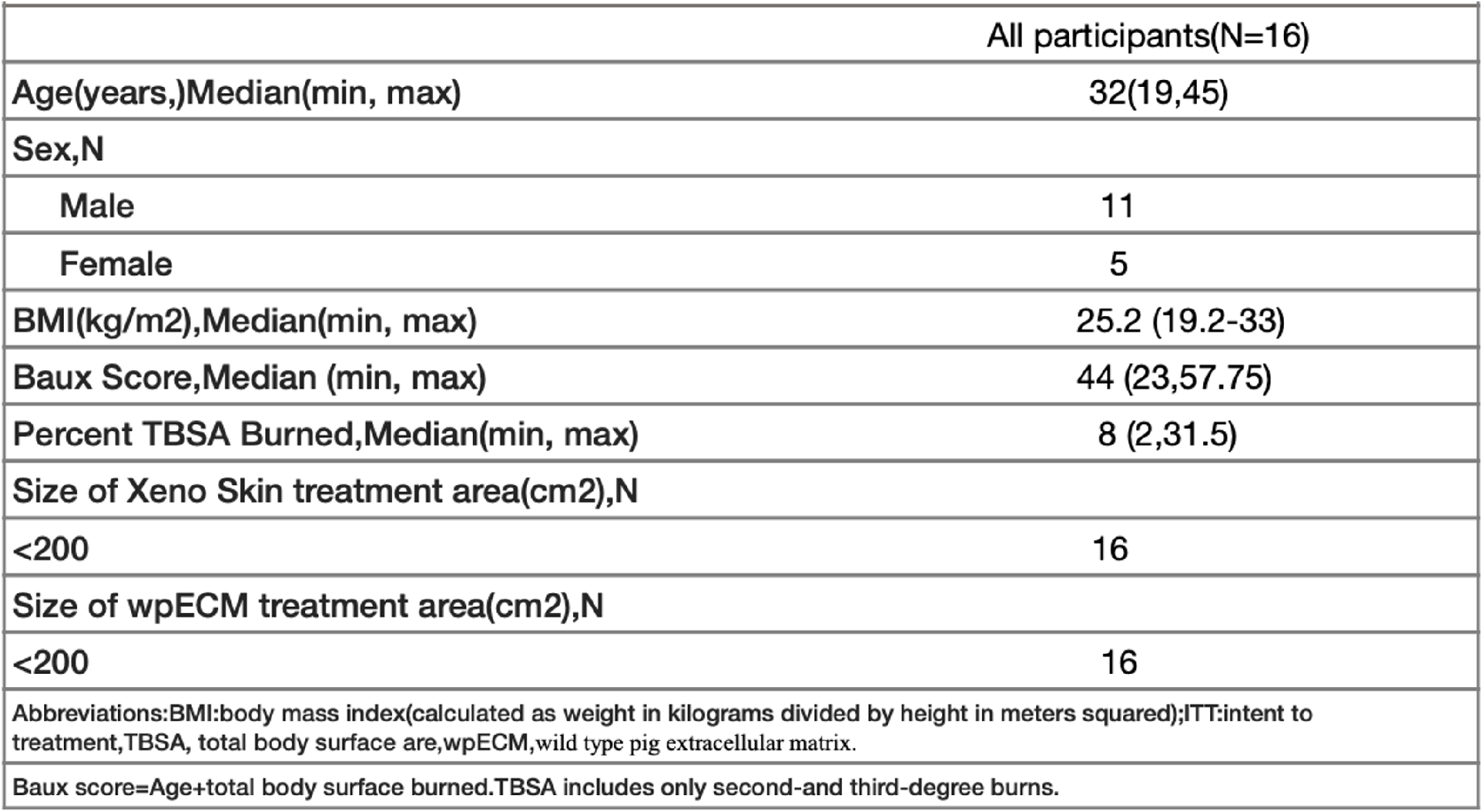
Summary of Demographics and Baseline Characteristics of ITT Population

### Safety

After the 4 weeks follow-up, neither adverse events nor wound infection was reported in all participants. Their vital signs and laboratory parameters were normal and there was no evidence to support any zoonotic disease transmission including porcine endogenous retroviruses (PERVs).

### Efficacy

The Xeno X Skin grafts from SGGEP can provide a durable wound closure time window for burned patients. The Xeno X Skin grafts in all patients were vascularized and fully adherent almost instantly (Figure 2A, Figure3A) without any immunosuppression. These grafts gave a rapid roughly 100% closure of burn wounds area from the post-transplant day1 to the post-transplant day8(Figure 2A,Figure3B).On the post-transplant day 14 the Xeno X Skin grafts accomplished over 80% closure of burn wounds although the rejection showed up as expected (Figure2A).On the post-transplant day 22,there was still approximately 30% closure of wounds area(Figure3C).The pathological assays(HE staining) illustrated that on the post-transplant day 14 and day 21 the Xeno X Skin graft exhibited vascularized and showed a healthy, complete skin structure which was in line with gross anatomy appearance(Figure4A,4B).

**Figure 2.**
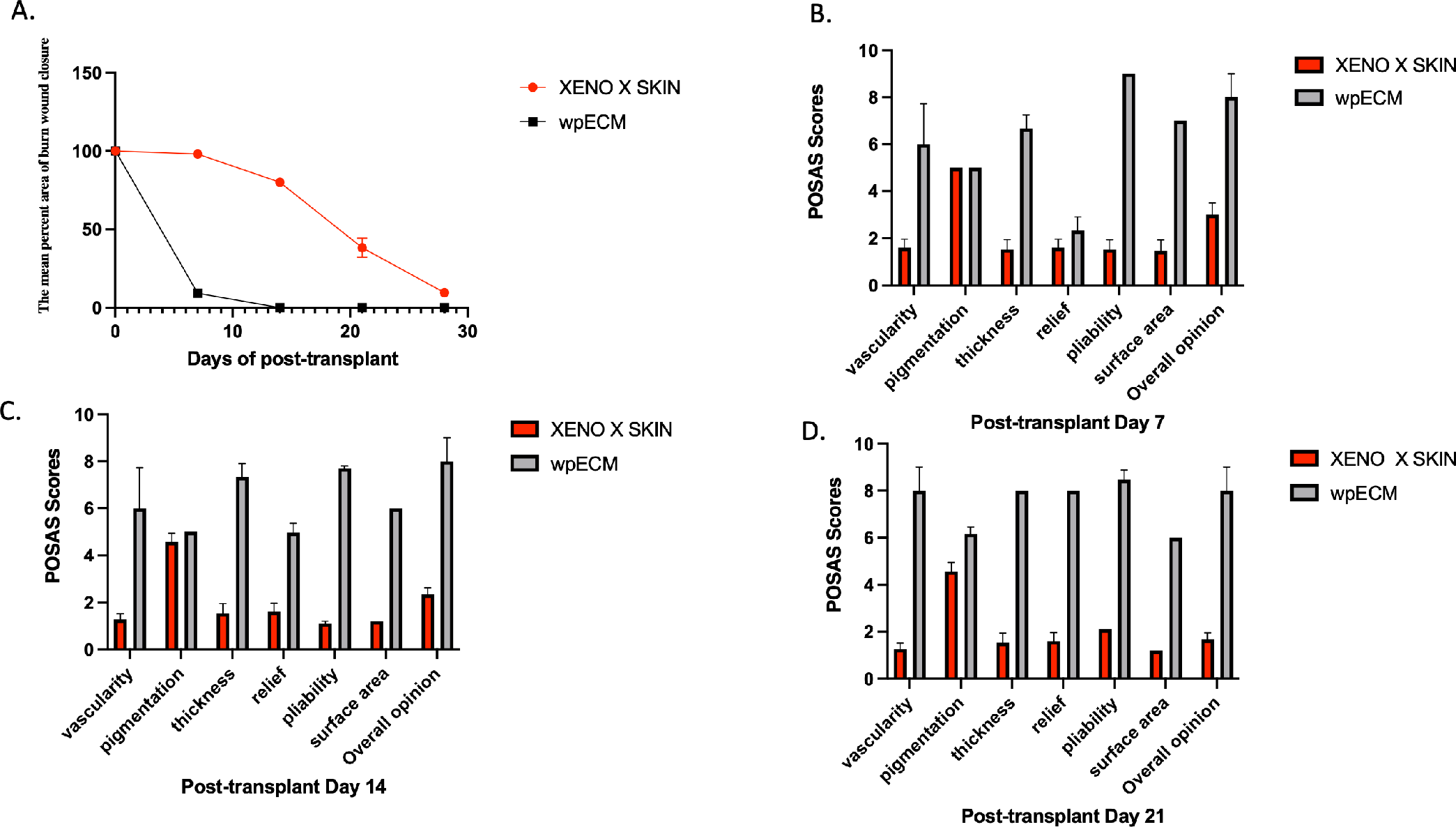
The percent of wound area achieved closure and wound POSAS observer scores of the skin xenotransplant and wild type pig extracellular matrix (wpECM) in a side-by-side, in-situ comparison without any immunosuppressor.(A)The mean of percent area of the wound closure with xeno X skin and wpECM over time.(B-C)POSAS scores the skin xenotransplant and wild type pig extracellular matrix (wpECM) over time.

**Figure 3.**
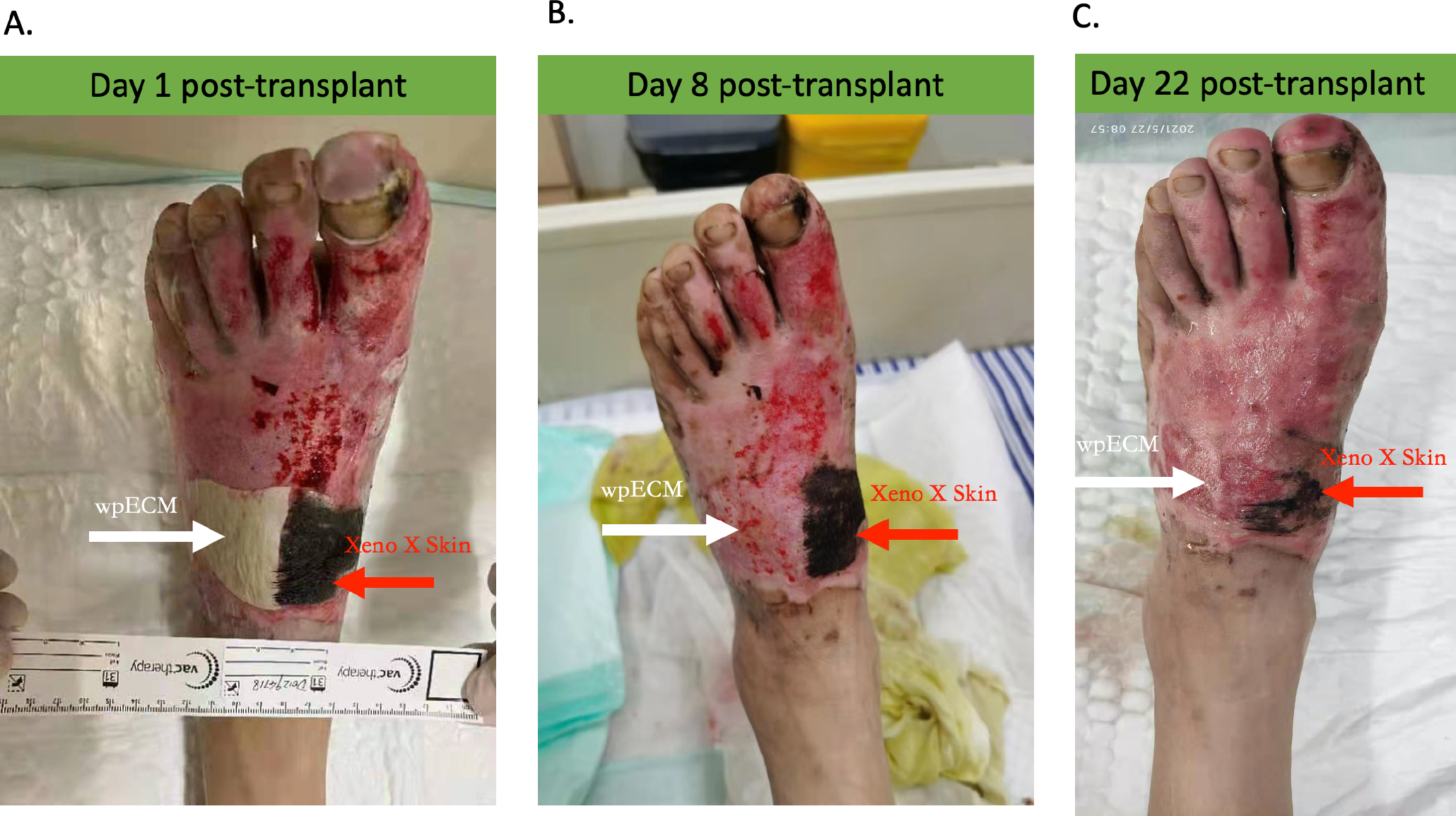
Photographic pictures of the typical skin xenotransplant and wild type pig extracellular matrix (wpECM) in a side-by-side, in-situ comparison. (A)Day 1 post-transplant. White arrow showed wpECM(wild pig extracellular matrix),red arrow showed Xeno X skin. Both the Xeno X skin and wpECM Xeno X skin were fully adherent, but only Xeno X skin was vascularized. (B) Day 8 post-transplant. White arrow showed wpECM (wild pig extracellular matrix),red arrow showed Xeno X skin.The wpECM was not adherent, but the Xeno X skin was still fully adherent and vascularized. (C) Day 22 post-transplant. White arrow showed wpECM(wild pig extracellular matrix),red arrow indicated Xeno X skin. The wpECM was not adherent; the Xeno X skin was still fully adherent and vascularized although 2/3 area was rejected.

**Figure 4.**
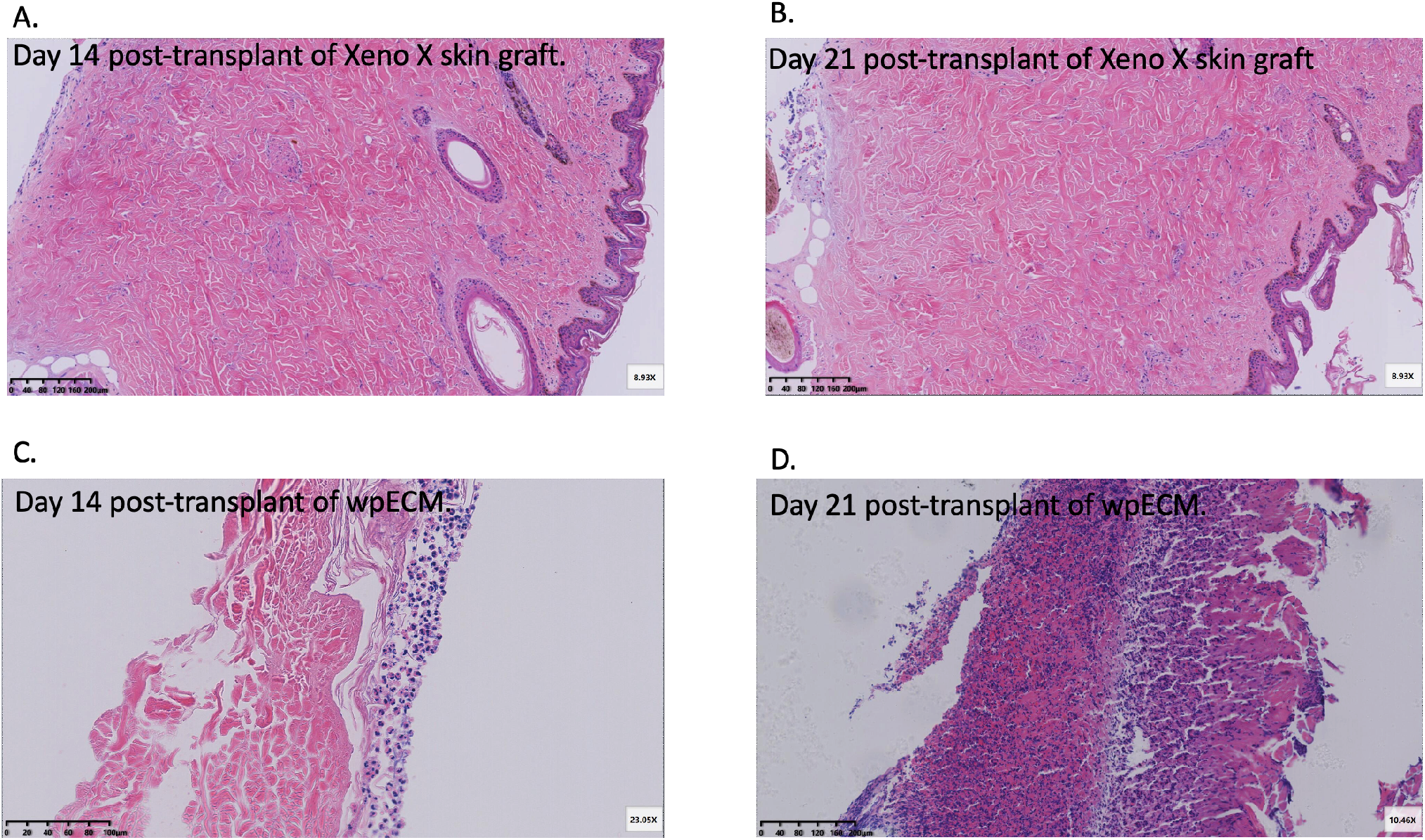
Pathological pictures of a typical skin xenotransplant and wild type pig extracellular matrix (wpECM) in a side-by-side, in-situ comparison. (A)Day 14 post-transplant of Xeno X skin graft. Consisting with the photographic picture, the graft bear normal skin structure and fully vascularized. (B) Day 21 post-transplant of Xeno X skin graft. Consisting with the photographic picture, the graft bear normal skin structure and fully vascularized. (C) Day 14 post-transplant of wpECM. In agreement with photographic pictures, the wpECM was not vascularize and adherent. (D) Day 21 post-transplant of wpECM. In agreement with photographic pictures, the wpECM was not vascularize and adherent.

In contrast, the wpECM grafts could adhere to wounds on post-transplant day 1 but they were never vascularized as anticipated and shed completely on the post-transplant day 8(Figure2A, Figure3B). The pathological assays (HE stains) demonstrated that there is not a complete skin structure on the post-transplant day 14 and day 21(Figure4C,4D).

The mean donor-site POSAS observer scores on the post-transplant day7, day14, day 21 for every category of vascularization, pigmentation, thickness, relief, pliability, and surface area are summarized in Figure 2B, 2C,2D. Scores for every POSAS category were lower for Xeno X skin wound sites comparing with wpECM donor sites, revealing they were much more like normal skins.

Strikingly, the Xeno X Skin grafts were recovered from over 1 year cryopreservation and demonstrated a healthy histopathological structure and good performance as the sustainable temporary wound coverage. These results are in compliance with the other report in which the pig skin grafts were were recovered from 7 years of storage^10^.

Together, our X prospective cohort study of Xeno X skin graft in patients with thermal burns exhibited enhanced immunological compatibility with human, well consistent with our preclinical research in non-human primates. These findings provided direct evidence that organ xenotransplant has become a clinical reality.

## Discussion

Recent study in large animal models witnessed the progress in the creation of viable, and functional skin xenografts with clinically oriented shelf-lives to upgrade the treatment of severe burn patientss^10,11^.These progress lay down the basis for the first FDA-approved phrase I clinical trial of a live skin xenograft for the burned patients(ClinicalTrials.gov Identifier:NCT03695939).But, so far there is no prospective cohort research data reported.

To our knowledge, the present prospective cohort study is the first one to assess the safety and efficacy of the cryopreserved SGGEP’s skin grafts (Xeno X skin), putting Xeno X skin graft and wild type pig extracellular matrix (wpECM) in a side-by-side, in-situ comparison, in the absence of any immunosuppressor. Our findings demonstrated that overall, the safety of Xeno X skin is reassuring, no advent events or unexpected patterns of concern were recognized. Comparing to wpECM the Xeno X skin grafts were vascularized and fully adhered to the wound bed. These skin xenografts could achieve at least 25 days graft survival without immunosuppressive drugs,which means the Xeno X skin could produce enough life-saving time window for severe and intensive burned patients.

In comparison, the allograft and single gene modified pig xenograft could only provide 8 days’ wound closure time in patients and 13 day’s wound closure time in non-human primates respectively^6-8^. On top of that, although the human cadaveric allograft remains the mainstream of burn management, cadaveric skin, like other organs, is vulnerable to global shortages, high cost and infectious disease concerns.

Therefore, our unprecedented work demonstrates that the Xeno X skin could be an effective treatment for severe burns, particularly when the human cadaveric skin is in short supply.

The clinical value of our work is dramatically strengthened by the facts that Xeno X skin could be preserved, and long-term storage could be achieved without significant loss of viability. In our trial the xenografts showed viable skin function, as well as the healthy and complete skin structure. These findings indicated that vital porcine skin grafts that can be successfully stored and shipped worldwide, which in turn will improve emergency preparation for an unexpected disaster or war.

Initially,there was a theoretical concern about the porcine endogenous retroviruses (PERVs) ^12,13^.However,porcine tissue has been applied for decades as temporary closure in burn treatment without any sign of zoonotic transmission^12^. Recently, genetically engineered PERV knockout animals have been established; however, the threat of off target mutations has been regarded to be greater than that of actual zoonotic transmission^12,13^. More importantly the PERV knockout will sacrifice the valuable and limited numbers of gene modifications in pigs since excessive gene edition will be detrimental to pig organs’ function and it may will lead to aborting in the surrogate pigs^14^.

The key limitation of the data is the shortage of a direct a side-by-side, in-situ comparison between Xeno X skin graft and deceased skin graft. Another limitation lies in the short durationof the flow-up. The trial is ongoing, inclusion of human cadaveric graft in the comparison study, as well as and a 2 years follow-up are planned.

The data presented in this study have significance beyond the performance of this Xeno X skin graft candidate. The skin is not only the largest organ of the body but also it is the highly immunogenetic (the skin harbors a wide collection of immune cells^15,16^). Our results illustrate that long immune tolerance of Xeno X skin in human without immunosuppression could be achieved through comprehensive gene modifications of pigs.

Therefore, other clinical organ xenotransplants such as cornea, kidney, heart, liver xenotransplantation would be accomplished successfully through the same genetic modification strategy in this study and related trials are also planned.

Additionally, our genome editing technologies in pigs could have other application and benefit. For instance, our work has tremendous effects on upgrading current and future pig biomedical models, while humanized (gene edited) pig models can be utilized in preclinical research^17^. Besides, since the pig is also an important food source, people could apply these genome editing technologies for generating the specific food for certain patients.For example, the U.S FDA has recently authorized the GalSafe gene edited pigs^18,19^as the food to help people who are under the effects of tick bite–induced allergic reactions to the sugar, galactose, which is also known as galactose-α1,3-galactose (α-Gal) syndrome (Figure 5).

**Figure 5.**
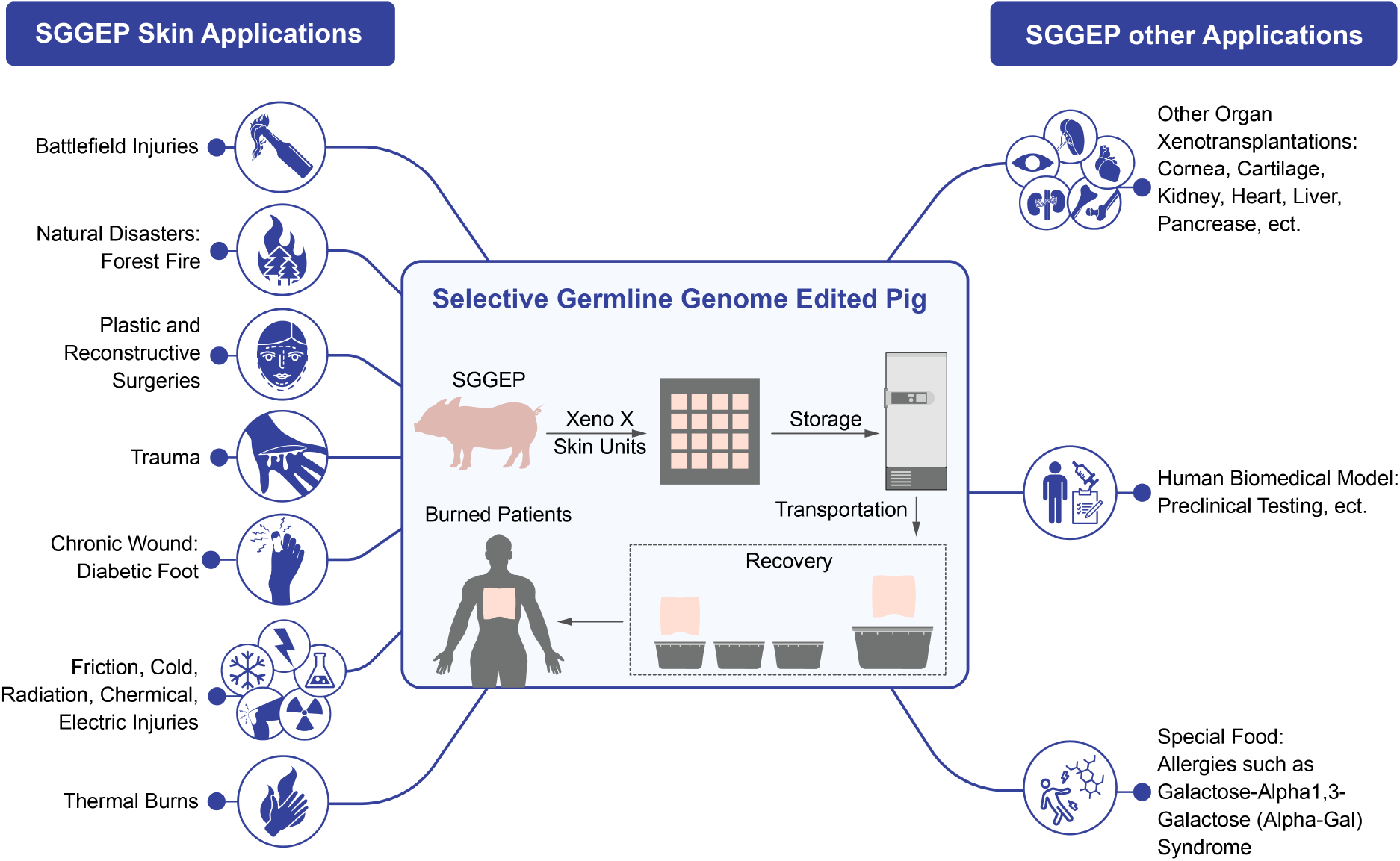
Applications of the selective germline genome gene edited pig

## Data Availability

All data produced in the present study are available upon reasonable request to the authors

## Acknowledgements

The authors thank the study participants and the authors thank Yuting Yang,Yuanyuan Zhang and Shi Chen for administrative support.

